# Longitudinal changes in the cortico-reticulospinal tract associated with high-intensity locomotor training in chronic stroke

**DOI:** 10.64898/2026.05.12.26353042

**Authors:** Jolene Foster, Oluwole O. Awosika, Pierce Boyne

**Affiliations:** Department of Rehabilitation, Exercise, and Nutrition Sciences, College of Allied Health Sciences, University of Cincinnati, OH, USA; Department of Neurology and Rehabilitation Medicine, College of Medicine, University of Cincinnati, OH, USA

**Keywords:** brain, gait, locomotion, stroke, contralesional, compensation

## Abstract

**Introduction:** High-intensity locomotor training (HIT) is recommended for improving walking capacity, but treatment responses are variable. Understanding the brain changes underlying responsiveness to training could provide insight into this variability. Emerging evidence suggests upregulation of the contralesional cortico-reticulospinal tract (CRST) may contribute to walking function after stroke. However, it is unclear whether CRST upregulation is supportive or maladaptive, and no studies have examined CRST changes after HIT. This study investigated how CRST and corticospinal tract (CST) strength and laterality reorganize, and their relationship with walking capacity after locomotor HIT.

**Methods:** Ten participants with chronic stroke completed a 4-week no-intervention control phase then 4-weeks of HIT. Diffusion MRI and 6-minute walk distance were obtained at weeks 0, 4, and 8. Analysis tested changes in ipsilesional and contralesional CRST and CST strength and laterality. Associations between changes in tract laterality and walking capacity were examined.

**Results:** During the treatment phase (vs. the control phase), there were significantly greater increases in contralesional CRST strength (1.02 SD [95% CI: 0.25, 1.79]), contralesional CRST laterality (4.44 [2.15, 6.72]), and 6-minute walk distance (33 meters [17, 50]). Walking capacity improvements were associated with changes in CRST laterality (r = 0.77, p = 0.01), but not CST laterality (r = -0.01, p = 0.98).

**Discussion:** Following HIT, increases in contralesional CRST strength and laterality were observed. CRST laterality changes were strongly associated with walking improvements, suggesting a possible supportive role of contralesional CRST in mediating training-related improvements in walking function after stroke.

**Clinical trial registration:** ClinicalTrials.gov Identifier: NCT02858349, https://clinicaltrials.gov/study/NCT02858349?term=NCT02858349&rank=1

## Introduction

Persistent walking impairments and decreased walking capacity remain leading causes of long-term disability in chronic stroke.^1^ Clinical practice guidelines for stroke rehabilitation recommend moderate- to high-intensity locomotor training to improve walking outcomes.^2^ This approach has demonstrated clinically meaningful improvements in walking capacity, especially with high-intensity locomotor training (HIT),^3,4^ but treatment responses remain variable.^3,4^

Improved understanding of the brain processes driving response to HIT may provide insight into variability of treatment response and guide future interventions. For example, non-invasive brain stimulation is a potential adjunct to gait training to enhance neuroplasticity and improve walking function after stroke.^5,6^ Effective integration of such approaches requires an understanding of how HIT influences reorganization of brain motor pathways involved in walking after stroke to identify candidate neural targets.^5–9^ However, the brain motor pathway changes induced by HIT remain unknown.

In studies examining the neural correlates of walking function after stroke, the corticospinal tract (CST) has been the primary focus.^10–13^ However, the CST alone has not fully explained gait function. For example, individuals with near to complete CST disruption are able to regain independent walking,^10–13^ suggesting that alternative descending motor pathways likely contribute to walking recovery.

One such pathway is the contralesional cortico-reticulospinal tract (CRST), which has been shown to be upregulated after stroke,^10,14–16^ thus shifting the balance of ipsilesional vs. contralesional motor pathway strength further towards contralesional lateralization. The contralesional CRST primarily runs ipsilaterally to influence the hemiparetic limbs, providing an anatomical substrate for contralesional compensation to reach the paretic limbs.^17,18^ The functional significance of contralesional CRST upregulation in walking function remains unclear, as prior cross-sectional studies have associated contralesional CRST upregulation with both better^10^ and worse^14^ motor outcomes. To our knowledge, no studies have examined longitudinal CRST changes in response to gait training in individuals with chronic stroke.

The aim of this study was to: (1) examine longitudinal changes in CRST strength and laterality; and (2) investigate associations between changes in CRST laterality and training-related changes in walking capacity in patients with chronic stroke. We also tested CST changes for comparison. We hypothesized that (1) CRST laterality would shift toward the contralesional hemisphere following 4 weeks of HIT; and (2) changes in CRST laterality would be significantly associated with changes in walking capacity.

## Materials and Methods

The study providing data for this analysis was approved by institutional review boards, preregistered on ClinicalTrials.gov (NCT02858349) and performed in a cardiovascular stress laboratory, MRI research center, and rehabilitation research laboratory from July 2016 to December 2017. Some of the methods and data from this study have been previously described in manuscripts addressing different aims from the current report.^16,19–21^

### Study Design

After screening and baseline assessment,^19,21^ each participant had a 4-week control phase with no-intervention, followed by a 4-week treatment phase with 12 sessions of locomotor high-intensity interval training performed 3x/week. Blinded gait testing and brain MRI were collected for each participant at baseline (week 0), after the 4-week control (week 4), and again after the 4-week treatment phase (week 8).

### Participants

Ten participants with chronic stroke were recruited from the community and provided written informed consent. Inclusion criteria were: age 30–90 years; unilateral stroke in middle cerebral artery territory experienced >6 months prior to enrollment; walking speed <1.0 meters per second on the 10 meter walk test ^22^; and able to walk 10 meters over ground with assistive devices as needed without physical assistance. Exclusion criteria were: MRI incompatibility; inability to perform mental imagery ^23^; evidence of significant arrhythmia or myocardial ischemia on treadmill electrocardiogram stress test, or significant baseline electrocardiogram abnormalities that would make an exercise electrocardiogram uninterpretable^24^; recent cardiopulmonary hospitalization; unable to communicate with investigators or correctly answer consent comprehension questions; significant ataxia or neglect (National Institutes of Health Stroke Scale item score >1)^25^; severe lower extremity hypertonia (Ashworth >2) ^26^; recent drug or alcohol abuse or significant mental illness; major post-stroke depression (Patient Health Questionnaire-9 ≥ 10)^27^ in the absence of management of the depression by a health care provider^28^; participating in physical therapy or another interventional research study; recent paretic lower extremity botulinum toxin injection; concurrent progressive neurologic disorder or other major conditions that would limit capacity for improvement; and pregnancy.

### Locomotor High-Intensity Interval Training Treatment Protocol

This study used the strategy of locomotor high-intensity interval training, where bursts of effort alternate with recovery periods during walking. Each session alternated between short and long-interval HIT sessions.^29^ Short-interval HIT sessions involved alternating 30 second bursts at maximum safe speed with 30–60 seconds resting recovery periods.^29,30^ Burst speed was progressed as able throughout the session to maintain constant challenge. Long-interval HIT sessions involved alternating 3–4 minute bursts at a target heart rate of 90% peak heart rate with 2–3 minute active recovery periods at a target of 70% peak heart rate.^29,30^ Peak heart rate was the highest heart rate achieved during graded exercises stress testing prior to enrollment.^19,24^ Speed was continually adjusted to maintain the target heart rate.

Each locomotor high-intensity interval session included a 3-minute warm up (overground walking at ∼40% heart rate reserve) followed by, 10-minutes of overground HIT, 20-minutes of treadmill HIT, and another 10-minutes of overground HIT, and a 2-min cool down (overground walking at ∼40% heart rate reserve). Heart rate reserve beats per minute targets were calculated by: (peak heart rate – resting heart rate) × % heart rate reserve target + resting heart rate).^24^ Resting heart rate was the resting heart rate before each training session.^19,24^ Participants utilized baseline orthotic and assistive devices for all walking and wore a fall protection harness during treadmill training. Physical assistance was provided only when needed to prevent a fall or injury. During overground training, participants walked back and forth in a corridor, using visual feedback for distance covered and verbal encouragement to maximize speed. Assistive device and/or walking pattern were progressed as able to achieve faster speeds.

### Walking Capacity Assessment

Walking capacity was measured by the 6-minute walk distance. During the test, the participant is instructed to walk as far as possible in 6 minutes using a standardized course.^31^ The distance walked provides a reliable and valid measure of walking capacity and is associated with community ambulation after stroke.^32,33^ A blinded rater administered the test.

### MRI Data Acquisition

A 3.0T Philips Ingenia MRI system was used. T1-weighted brain images were acquired at 1mm isotropic resolution with the following parameters: TR, 8.1ms; TE, 3.7ms; flip angle, 8^0^; SENSE factor 2. Diffusion weighted brain images were acquired at 2mm isotropic resolution with the following parameters: TR, 7.1s; TE, 92ms; flip angle, 90^0^; 61 directions at b=1000 s/mm^2^; 7 b=0 volumes; SENSE factor 3.

### MRI Data Preprocessing

For the T1 images, FSL software^34^ was used for bias field correction, tissue type segmentation and non-linear registration to the MNI152 template, using a lesion mask to improve registration for participants with stroke by temporarily filling the lesion with MNI template voxels.^35^ Diffusion MRI preprocessing included eddy current correction, motion correction, and outlier volume replacement using FSL’s “eddy” tool.^34,36,37^ Alignment to the T1 image with boundary-based registration was performed using the T1 white matter segmentation.^34,38^ Diffusion data were reconstructed with generalized Q sampling imaging using DSI Studio (https://dsi-studio.labsolver.org/) with a diffusion length ratio of 1.7, to calculate quantitative anisotropy in each brain voxel.^39,40^ Diffusion anisotropy measures such as fractional anisotropy and quantitative anisotropy are common markers of post-stroke tract microstructural integrity,^41^ neural reorganization,^42^ and motor tract projection strength.^41^ Compared to fractional anisotropy, quantitative anisotropy is a more specific anisotropy measure that provides improved resolution in areas of multiple fiber directions and is less sensitive to partial volume effects and edema.^41^ Lower anisotropy is a marker of motor tract damage and decreased tract strength, while higher anisotropy is a marker of neural reorganization and increased tract strength (i.e. tract upregulation/compensation).^42–45^

### Tract Strength Measurement

The following steps were performed for calculating the motor tract strength of each motor tract (ipsilesional CST and CRST; contralesional CST and CRST):^16^

1. Quantitative anisotropy values were normalized (nQA) to reduce the impact of scanner variability by dividing the quantitative anisotropy value in each voxel by the mean quantitative anisotropy within the ventricular cerebrospinal fluid from each specific scan.^46^
2. The nQA map was registered to a standard brain coordinate space (MNI152 6^th^ generation template distributed within ‘FSL’ software),^34^ where local nQA maxima were then projected onto a core white matter “skeleton” (distributed within ‘FSL’)^47^ to further improve registration and account for anatomical variability.^48,49^
3. Mean nQA values from each motor tract (ipsilesional CST and CRST; contralesional CST and CRST) were calculated, with voxel averaging weighted by the number of normative tract streamlines in each voxel, within the internal capsule region (z = -5 to 12 mm) of each tract.^18,45,50^
4. The nQA measurement for each tract was adjusted for mean global cerebral white matter nQA to more specifically quantify the strength of each tract, independent from global white matter nQA. This was done by modeling the relationship between global nQA (independent variable) and tract-specific nQA (dependent variable), averaged across left and right tracts. Only control participant data were used for this modeling, so that we could use it to calculate how the tracts of stroke survivors differed from the expected normative value for someone with the same global QA, but without tract damage.^16^
5. Tract nQA values were then converted to z-scores using control data to improve interpretability. The control mean and standard deviation values used to calculate the z-scores are available in a prior manuscript.^16^

### Tract Laterality

Tract strength laterality was used as a relative measure of contralesional vs. ipsilesional motor tract strength.^44,51^ Laterality was calculated by: (contralesional tract strength – ipsilesional tract strength) / (contralesional tract strength + ipsilesional tract strength) x 100. Values range from -100 (completely ipsilesional) to +100 (completely contralesional), with 0 representing complete symmetry.^51^

### Sample Size Calculation

The target sample size of 10 participants was calculated for different aims than those reported here.^20^ That sample size provides 80% estimated power to detect: 1) a within-group Cohen’s d effect size as small as 1.00; 2) a between-group effect size as small as 1.32; and 3) a correlation as low as 0.58. These calculations were based on a two-sided significance level of 0.05 and were performed with the R package ‘pwr’.^20^

### Data Analysis

To assess how tract changes related to walking capacity changes, we performed two analyses. First, linear mixed-effects models were used to assess longitudinal changes in tract strength and laterality of the CRST and CST, with week modeled as a categorical variable. Participant-specific random effects for week were used to account for within-subject correlation across repeated measures. Models were fit using the R package ‘nlme’. For participant *i* at week *j*, the tract strength or laterality (Y*_ij_*) was modeled as:

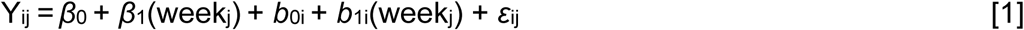

where *β*_0_ and *β*_1_ are the fixed intercept and week effects, *b*_0i_ and *b*_1i_ are participant-specific random effects for week, and *ε*_ij_ is the residual error.

Second, within-participant associations between CRST laterality change and 6-minute walk distance change were assessed using repeated-measures correlations (R package ‘rmcorr’)^52^ after adjusting each tract change and 6-minute walk distance change for changes in the other tract. To obtain these associations in unstandardized units (i.e. the expected difference in 6-minute walk distance change for each 1 standard deviation higher change in tract z-score), additional linear mixed-effects models similarly examined the independent contributions of CRST and CST laterality change to 6-minute walk distance change, with participant-specific random effects to account for repeated measures. For participant *i* at phase *j* (treatment or control phase), 6-minute walk distance change was modeled as:

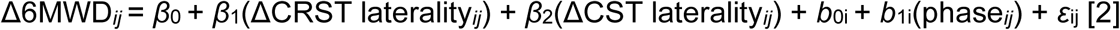

where *β*_0_- *β*_2_ are fixed effects, *b*_0i_ and *b*_1i_ are participant-specific random effects, and *ε*_ij_ is the residual error. All analyses were conducted using R version 4.4.1. The STROBE reporting guidelines^53^ were used to draft this manuscript, and the STROBE reporting checklist^54^ was used when editing and is included in supplement A.

## Results

The target sample size of 10 participants (Table 1) was achieved and there were no missing data. The strength and laterality measurements across weeks 0, 4, and 8 foreach motor tract examined are reported in Table 2.

**Table 1.** Participant characteristics.

|  |  |
| --- | --- |
| Age (years) | 59.8 ± 6.78 |
| Gender, N (%) |  |
| Female | 4 (40.0%) |
| Male | 6 (60.0%) |
| Body Mass Index (kg/m<sup>2</sup>) | 30.2 ± 4.24 |
| Comorbid conditions, N (%) |  |
| Diabetes mellitus | 1 (10%) |
| Hypertension | 6 (60%) |
| History of percutaneous coronary intervention | 2 (20%) |
| History of myocardial infarction | 1 (10%) |
| History of seizure | 2 (20%) |
| Years since stroke | 2.4 ± 1.7 |
| Type of stroke, N (%) |  |
| Hemorrhagic | 1 (10.0%) |
| Ischemic | 9 (90.0%) |
| Lesion side, N (%) |  |
| Left | 5 (50.0%) |
| Right | 5 (50.0%) |
| Lesion volume, mL | 122 ± 105 |
| PHQ-9 score | 3.1 ± 3.1 |
| Assistive device, N (%) |  |
| Hemiwalker | 1 (10.0%) |
| Wide based quad cane | 4 (40.0%) |
| Narrow based quad cane | 2 (20.0%) |
| None | 3 (30.0%) |
| Orthotics, N (%) |  |
| AFO | 8 (80.0%) |
| None | 2 (20.0%) |
| Comfortable gait speed (m/s) | 0.41 ± 0.33 |
| Six-minute walk distance (m) | 156 ± 147 |

Values are mean ± SD or N (%). Abbreviations: NA, not applicable. PHQ-9, Patient Health Questionnaire-9 total score.

**Table 2.** Tract strength, tract laterality and walking capacity by week.

|  | Control Phase |  | Training Phase |
| --- | --- | --- | --- |
|  | Week 0 | Week 4 | Week 8 |
| **Tract strength z-scores (SD)** |  |  |  |
| iCRST | -8.59 (3.18) | -8.37 (3.49) | -8.67 (3.16) |
| cCRST | 1.52 (1.65) | 0.96 (1.31) | 1.42 (1.41) |
| iCST | -9.78 (3.08) | -9.89 (3.12) | -9.37 (3.23) |
| cCST | 2.87 (3.14) | 2.22 (2.40) | 2.58 (2.56) |
| **Tract strength laterality** |  |  |  |
| CRST laterality | 22.1 (12.4) | 19.9 (12.8) | 22.2 (11.4) |
| CST laterality | 34.6 (12.1) | 33.9 (12.0) | 32.5 (12.5) |
| **Walking Capacity** |  |  |  |
| 6-Minute Walk Distance (m) | 156 (147) | 154 (156) | 186 (173) |

Mean standardized tract strength (SD, standard deviations), laterality z-scores (SD), and walking capacity values (meters, SD) by study week. Tract measures were standardized using a reference distribution prior to analysis. Abbreviations: iCRST, ipsilesional cortico-reticulospinal tract; cCRST, contralesional cortico-reticulospinal tract; CRST, cortico-reticulospinal tract; iCST, ipsilesional corticospinal tract; cCST, contralesional corticospinal tract; CST, corticospinal tract.

There was an increase in contralesional CRST tract strength during the training phase which was significantly greater than the control phase change (1.02 SD [95% CI: 0.25, 1.79]), whereas no significant changes were detected in the remaining motor tracts assessed. CRST laterality demonstrated a further contralesional shift during training (2.27 [0.88, 3.66]), and this change was significantly greater than the control phase change by 4.44 [2.15, 6.72]. In contrast, the CST shifted non-significantly toward the ipsilesional side during the training phase (-1.35, [-2.98, 0.28]) and had a non-significant training-versus-control phase difference (-0.68 [-3.92, 2.56]). In parallel, 6-minute walk distance increased during the training phase (31 meters [17, 46]), exceeding the control phase by 33 meters [17, 50] (Table 3).

**Table 3.** Changes in tract strength, tract laterality, and walking capacity.

|  | **Control phase**<br>**change**<br>**(4WK – WK0)** | **Training phase**<br>**change**<br>**(8WK – 4WK)** | **Training – control**<br>**phase change** |
| --- | --- | --- | --- |
| **Tract strength z-scores** |  |  |  |
| iCRST | 0.22 [-0.35, 0.79] | -0.30 [-0.84, 0.24] | -0.52 [-1.49, 0.45] |
| cCRST | -0.56 [-1.12, 0.01] | 0.46 [0.12, 0.81] | 1.02 [0.25, 1.79] |
| iCST | -0.11 [-0.53, 0.30] | 0.52 [-0.02, 1.06] | 0.63 [-0.01, 1.28] |
| cCST | -0.65 [-1.75, 0.45] | 0.36 [-0.36, 1.08] | 1.01 [-0.27, 2.29] |
| **Tract Laterality** |  |  |  |
| CRST laterality | -2.16 [-3.58, -0.75] | 2.27 [0.88, 3.66] | 4.44 [2.15, 6.72] |
| CST laterality | -0.67 [-2.70, 1.36] | -1.35 [-2.98, 0.28] | -0.68 [-3.92, 2.56] |
| **Walking Capacity** |  |  |  |
| 6-Minute walk<br>distance (m) | -2 [-12, 8] | 31 [17, 46] | 33 [17, 50] |

Values are contrasts (estimate [95% CI]) from linear mixed-effects models including participant-specific random intercepts and slopes. Abbreviations: WK0, week 0; 4WK, week 4; 8WK, week 8; iCRST, ipsilesional cortico-reticulospinal tract; cCRST, contralesional cortico-reticulospinal tract; CRST, cortico-reticulospinal tract; iCST, ipsilesional corticospinal tract; cCST, contralesional corticospinal tract; CST, corticospinal tract.

When investigating associations between longitudinal changes in tract laterality and walking capacity, repeated-measures correlation analysis showed that walking capacity change was positively associated with change in CRST laterality (adjusted for CST laterality) (r = 0.77, p = 0.01) but not change in CST laterality (adjusted for CRST laterality) (r = −0.01, p = 0.95). Likewise, linear mixed-effects modeling demonstrated that change in 6-minute walk distance was independently associated with change in CRST laterality (5.85 meters/SD [2.54, 9.16]), but not CST laterality (-0.18 meters/SD [-3.77, 3.42]) (Table 4).

**Table 4.** Associations between tract laterality changes and 6-minute walk distance change.

| Variable | Estimate | Std. Error | t value | Pr(>\|t\|) |
| --- | --- | --- | --- | --- |
| (Intercept) | 13.62 | 3.88 | 3.51 | 0.01 |
| CRST laterality change | 5.85 | 1.43 | 4.08 | 0.00 |
| CST laterality change | -0.18 | 1.56 | -0.11 | 0.91 |

Random effects: SD(intercept) = 14.64, SD(phase) = 25.46, Residual SD = 7.99
Abbreviations: CRST, cortico-reticulospinal tract; CST, corticospinal tract.

## Discussion

This study examined longitudinal changes in the CRST following locomotor high-intensity interval training in individuals with chronic stroke, while also assessing CST changes for comparison. At baseline, CRST and CST strength laterality were shifted towards the contralesional hemisphere (Table 2). During the training phase, participants demonstrated a significant increase in contralesional CRST strength, a further contralesional shift in CRST laterality, and a significant increase in 6-minute walk distance. No significant changes were observed in strength or laterality for the ipsilesional CRST, ipsilesional CST, or contralesional CST. These training phase increases were also significantly greater than control phase changes for contralesional CRST strength and laterality, and 6-minute walk distance (Table 3 and Figure 1). Changes in CRST laterality were strongly associated with changes in walking capacity, whereas CST laterality changes were not (Figure 2). These findings suggest that upregulation of the contralesional CRST in response to HIT may promote adaptive reorganization of the contralesional CRST, which may contribute to better walking function.

**Figure 1.**
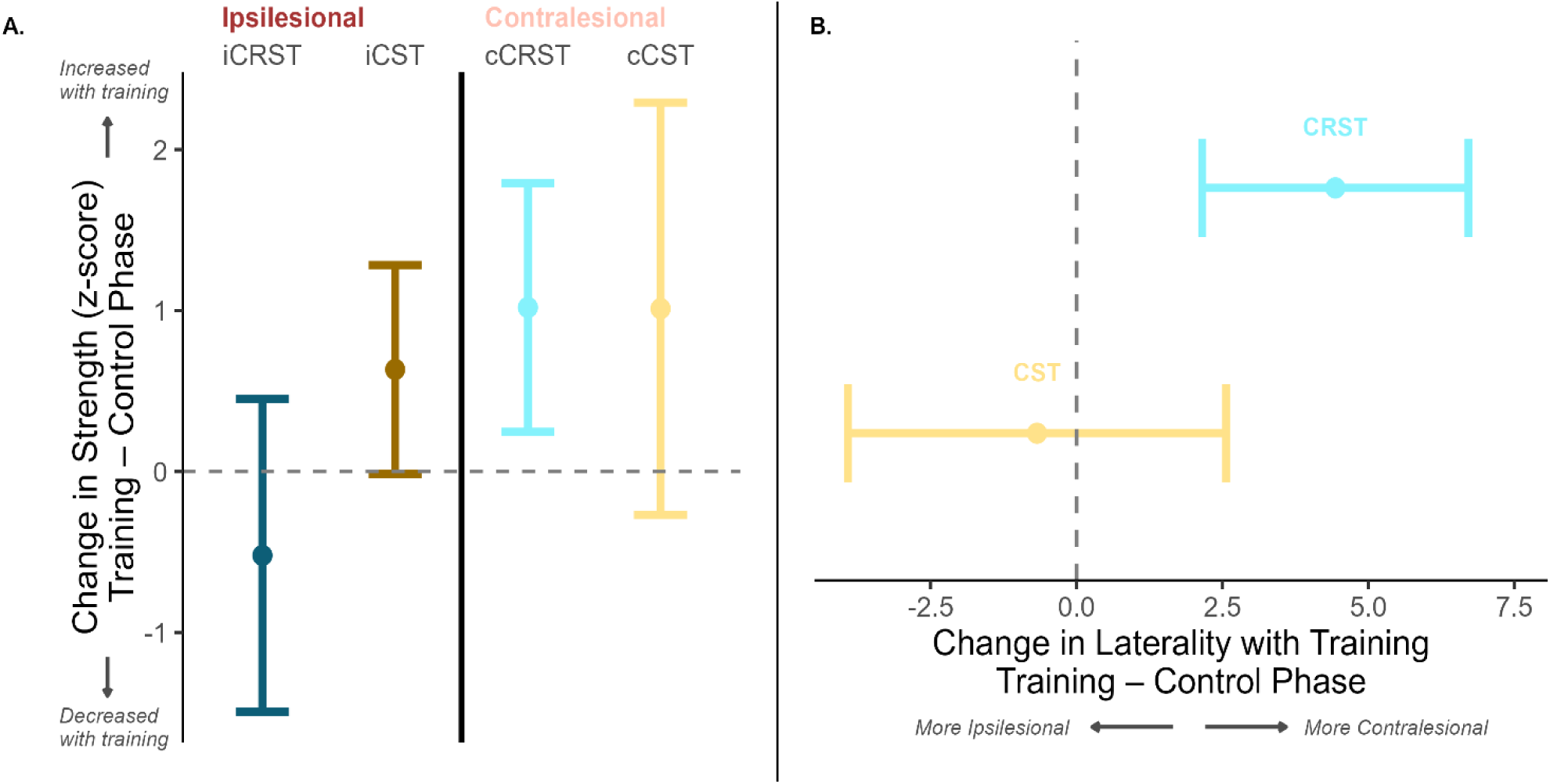
Motor tract changes after 4-weeks of HIT vs. 4-weeks of no-intervention. Points represent estimated mean differences in change derived from linear mixed-effects models, and error bars indicate 95% confidence intervals. (A) Training-minus-control phase changes in ipsilesional and contralesional cortico-reticulospinal (CRST) and corticospinal (CST) tract strength (z-scores). Positive values indicate increased tract strength during the training phase relative to the control phase. (B) Training-minus-control phase changes in tract laterality for the CRST and CST. Positive values indicate a shift toward the contralesional hemisphere, whereas negative values indicate a shift toward the ipsilesional hemisphere. The dashed vertical line denotes no difference in change between phases.

**Figure 2.**
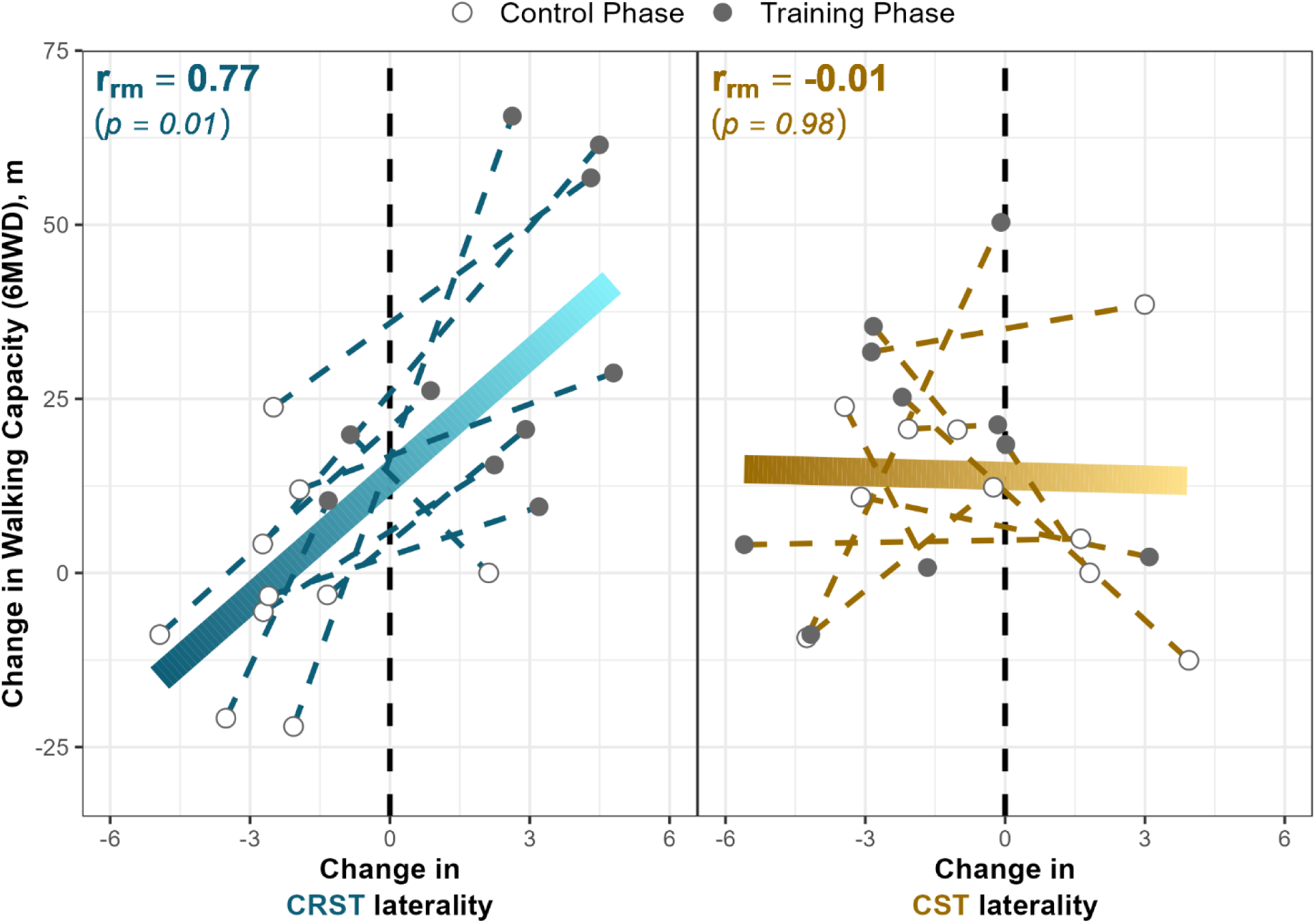
Training-induced shifts in CRST laterality correlated with walking gains. Associations between change in six-minute walk distance (6MWD) and cortico-reticulospinal tract (CRST) laterality and corticospinal tract (CST) laterality (adjusting for each other). Repeated-measures correlation (r_rm_) analyses were used to assess within-participant associations between training-minus-control phase changes in 6-minute walk distance (6MWD) and changes in tract laterality. Positive tract laterality changes indicate a shift toward the contralesional hemisphere, whereas negative changes indicate a shift toward the ipsilesional hemisphere.

Our finding that contralesional CRST strength increased alongside a contralesional shift in CRST laterality during training (Table 3 and Figure 1) is consistent with reorganization of this pathway in response to training. These neural changes occurred alongside a clinically meaningful increase in 6-minute walk distance of 33 [17, 50]) meters in the training phase (Table 3), with prior research showing a minimal clinically important difference of 14 to 30.5 meters.^55^ The increase in contralesional CRST strength also aligns with prior cross-sectional analyses suggesting the contralesional CRST is upregulated after stroke.^10,14–16^ Based on the current results, it now appears that HIT may further upregulate the contralesional CRST, thus lateralizing CRST strength further towards the contralesional hemisphere.

The strong association between changes in CRST laterality and walking capacity (r = 0.77) (Figure 2) suggests a functionally relevant role of the CRST in training-induced walking improvements after stroke. In contrast, CST strength and laterality did not change significantly and were not associated with walking improvements (Table 3 and Figure 2). These findings are consistent with prior studies suggesting the CST alone does not fully explain walking function after stroke.^10,12,56^ By extending prior cross-sectional evidence^11,18^ with a longitudinal design, the current findings also provide the strongest evidence to date supporting a possible compensatory role for the contralesional CRST after stroke.

In contrast to the training phase, an unexpected significant decrease in contralesional CRST laterality was observed during the control phase (Tables 2 and 3). Oddly, these neural changes were not accompanied by meaningful changes in walking capacity, as 6-minute walk distance remained relatively stable. One possible explanation for these findings is that low activity levels during the control phase may have contributed to reduced contralesional CRST strength, and that these neural changes may have preceded any potential walking changes we might have observed with a longer control phase. Future studies are needed to better understand these potential tract changes during the no-intervention control phase.

This study has potential implications for neuromodulation strategies aimed at enhancing walking function after stroke. Most non-invasive brain stimulation protocols aim to facilitate ipsilesional motor pathways and/or inhibit contralesional motor pathways.^5,8,9,57,58^ However, emerging evidence in both the upper extremity^8,9,57,59^ and lower extremity^5,10,16^ suggests that upregulation of contralesional motor pathways may support movement recovery in individuals with more severe stroke. Our previous work further demonstrated that the apparent negative association between contralesional CRST strength and walking capacity was confounded by the extent of ipsilesional motor tract damage, supporting the interpretation of contralesional CRST upregulation as an adaptive response.^16^ The present findings extend this work with a longitudinal and interventional design and provide further support for this interpretation. Future studies are needed to determine: (1) whether these findings vary based on stroke characteristics (e.g. severity of ipsilesional motor tract damage) and (2) whether non-invasive brain stimulation can augment the training-related tract changes occurring during HIT to further improve outcomes.

## Limitations

This study may have limited generalizability across the spectrum of stroke due to the relatively small sample size which consisted primarily of individuals with relatively severe stroke. Second, although a longitudinal analysis was performed, causal relationships between CRST changes and walking improvements cannot be definitively established and larger, randomized trials are needed. Additionally, the intervention period was relatively short (4-weeks) and prior research has shown 8- and 12-weeks of HIT training elicits greater gains in walking capacity.^3^ Thus, a longer intervention period may produce more pronounced brain changes. Finally, the study used a non-randomized cross-over design. A randomized cross-over design would require the absence of carryover effects into the subsequent no-intervention phase. However, evidence shows that walking capacity improvements after HIT are retained for up to 3-months, making carryover effects likely in the current design.^60^

## Conclusion

This study demonstrated that HIT appears to influence neural tract reorganization, with an increase in contralesional CRST strength and a contralesional shift in CRST laterality after 4-weeks of training. Changes in CRST laterality were associated with improvements in walking capacity change. These findings are consistent with prior work suggesting the contralesional CRST may be upregulated after stroke and provides longitudinal support that the CRST may have an important role in training-related improvements in walking function after stroke.

## Author contributions

JF: Methodology, Software, Formal analysis, Visualization, Writing – original draft, Writing-review & editing. OA: Methodology and Writing-review & editing. PB: Conceptualization, Data curation, Funding acquisition, Investigation, Methodology, Project administration, Resources, Software, Validation, Visualization, Writing – review & editing, Supervision. All authors contributed to the article and approved the submitted version

## Statements and Declarations

### Ethical considerations

This study received ethical approval by the University of Cincinnati IRB # 2016-1916

### Consent to participate

All participants provided written informed consent prior to enrolment in the study.

### Consent for publication

Not applicable

### Data Availability

The data that support the findings of this study are available from the corresponding author upon reasonable request.

